# Forecasting the CoViD–19 Diffusion in Italy and the Related Occupancy of Intensive Care Units

**DOI:** 10.1101/2020.03.30.20047894

**Authors:** Livio Fenga

**Affiliations:** Italian National Institute of Statistics, ISTAT, Rome, Italy 00184

**Keywords:** Autoregressive Moving Average Models, CoViD–19, Kalman filter, intensive care units, maximum entropy bootstrap

## Abstract

This paper provides a model–based method for the forecast of the total number of currently CoVoD-19 positive individuals and of the occupancy of the available Intensive Care Units in Italy. The predictions obtained – for a time horizon of 10 days starting from March 29th – will be provided at a national as well as at a more disaggregate levels, following a criterion based on the magnitude of the phenomenon. While the Regions which have been hit the most by the pandemic have been kept separated, the less affected ones have been aggregated into homogeneous macro-areas. Results show that – within the forecast period considered (March 29^*th*^ - April 7^*th*^) – all of the Italian regions will show a decreasing number of CoViD-19 positive people. Same for the number of people who will need to be hospitalized in a Intensive Care Unit (ICU). These estimates are valid under constancy of the Government’s current containment policies. In this scenario, Northern Regions will remain the most affected ones and no significant outbreak are foreseen in the southern regions.

## 1. Introduction

On March 19th, the death toll paid by Italy for the spread of the virus CoViD-19 amounted to 3405 deaths, the highest paid by a single country in the World. Despite an hard and relatively timely lock-down policy implemented by the Government, on March 26 this figure has risen to 8165 deaths.

In such an emergency situation, a reliable forecast method for the infection’s development is essential for policy and decision makers to design evidence-based policies and to implement fast actions to curb the spread of the infection. In particular, predicting the number of people currently tested positive for CoViD–19 (thereafter “positive cases”) could be useful to draw the epidemiological curve of the infection and therefore to predict its peak. Other than for this variable, the forecasting procedure presented in this paper is used to predict the future values of another crucial variable, i.e. the number of people needing hospitalization in a Intensive Care Unit (ICU). The Italian ICUs system is at the moment severely stressed due to the spread of the disease, therefore predictions of future ICUs demand could be fruitfully considered in the design and the implementation of operational schemes. The forecast horizon for both the variables is of 10-day starting from March 29th.

Since the Italian regions are affected in different extents by the CoViD-19, it has been decided to perform the forecasting exercise for the following geographical areas: Lombardia, Piedmont, Valle d’Aosta, Veneto, Friuli Venezia Giulia, Trentino Alto Adige, Lazio and Campania. The remaining Regions have been grouped in the following macro-areas: “Center” (Marche, Umbria and Toscana) and “South” (Abruzzo, Molise, Puglia Basilicata, Calabria, Sicilia and Sardegna). At least other two reasons justify such a break down:

1. the different starting times recorded for the lock-downs;
2. the Southern regions have been hit less severely and therefore, especially at the beginning of the observation period, show several zeroes or low numbers across the considered time span.

In essence, in this study the available official data have been employed in a three step procedure, i.e.:

1. data pre-processing, in which data anomalies are identified and corrected according to an approach of the type a Kalman filter;
2. univariate forecasting, based on a autoregressive moving average (ARMA) model for number of positive cases and ICU;
3. bootstrap–based generation of predicted values and confidence intervals.

## 2. The Data

This paper employs the data related to COVID-19, collected and regularly updated by the Italian National Institute of Health (an agency of the Italian Ministry of Health) and by the Italian Civil Protection Department. The whole data set is freely and publicly available in a comprehensive database, accessible on the Internet at the web address https://www.iss.it/. It collects crucial data related to all the persons tested for COVID-19 – from the outbreak of the pandemics (February the 24^*th*^) – and, in particular, it

1. is a collection of 21 data points – representing 19 Italian Regions plus the two autonomous provinces of Trento and Bolzano – for each day of infections;
2. considers crucial variables, such as positive cases, recovered cases, deaths, number of people hospitalized and number of people admitted to Intensive Cure Units (ICU).

As already pointed out, in the present study, the variables of interest are the number of people who have been:

1. tested positive for SARS-CoV-2 (in what follows denoted by the bold Latin letter **V**);
2. hospitalized in a ICU (which will be denoted by the bold Latin letter **U**).

It is worth to outlining how, according to the regulations issued by the Italian government, only the people showing moderate to severe symptoms, generally associated with the infection, or which have been in close proximity with at least one positive person, are tested. Therefore, the predictions obtained are to be referred to the sample, as no attempt have been made to carry out inferences procedures for variable estimation at the population level.

In order to correctly process the data, all the regions showing no positive cases at the beginning of the recording period and/or low values along the whole time span have been aggregated into macro areas. This has been done to *i*) give more meaningful results and *ii*) save degrees of freedom (which are always precious in short time series).

In details, the prediction exercise will be performed on the following Regions/macro areas:

A. Nothtern Regions
  1. Lombardia
  2. Piedmont
  3. Valle d Aosta
  4. Veneto
  5. Friuli Venezia Giulia
  6. Emilia Romagna
  7. Liguria
  8. Macro Area “Trentino Alto Adige” (Trento and Bolzano)
B. Center Regions
  1. Lazio
  2. Macro-area “Center” : Marche, Umbria and Toscana
C. Southern Regions
  1. Campania
  2. Macro-area “South” Abruzzo, Molise, Puglia, Basilicata, Calabria, Sicilia, Sardegna.

The north Italy Regions – presently the more severely affected by the pandemic – have been treated separately along with two other regions, i.e. Lazio and Campania, since their major cities – Rome and Naples – deserve special attention for the institutional role played and the population density exhibited. On the other hand, the Regions showing less worrying figures have been aggregated into macro areas according to their geolocation. The only exception is Valle d’Aosta, which has been left separated as no aggregation options could be found.

To simplify notation, for both the variables of interest *V* and *U*, the following convention is introduced:

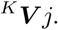

Here, the upper left superscript (denoted by the upper case Latin letter K) refers to the geographical areas (i.e. North, Center and South) whereas the subscript *j* is associated to the number the different regions or macroareas are codified with, as above detailed. For example, by the symbols ^*A*^***V***_6_ and ^*B*^***V***_2_ the number of positive cases for the Emilia Romagna region and the Center macro-area “Marche – Umbria – Toscana” are respectively identified.

## 3. Data Preprocessing

Missing data and other anomalies become the first challenge when designing predictive models, as statistical methods, in general, are designed and tested under the assumption of no missing observations (Moritz et al. (2015)).

Before delving into the details of the proposed procedure, a word of caution is needed since, unfortunately, a visual inspection of the database suggests the presence of a number of anomalous data both at a regional and country level. The detected anomalies might be associated to the biological samples collecting process and the testing procedures. In fact, the typical lab–workflow is governed by a set of rigid protocols which might be critically affected by factors such as the availability of manpower, swabs, reagents and other laboratory materials. In emergency situations, such a workflow can be disrupted and temporal inconsistency might appear as a result. For example, a set of samples might be delivered to a laboratory with longer than usual delays with respect to the time of collection or a given lab can only complete a screening process for a certain number of samples. In both the cases, one day (or more) shift in the release of the lab results can be reasonably expected. A further source of anomalies is represented by the data entry and data editing processes, carried out in working environment necessarily affected by the risk of contagious and under rigid deadlines.

An example of such anomalous data is given in Figure 1, where the series ^1^***V***_1,*t*_ (Lombardia) is depicted. Here, some data points showing values inconsistent with the overall pattern are clearly noticeable. Given the (very small) available sample size, the relative weight of such data is almost surely not negligible and can introduce severe distortions in the model parameter inference procedures and thus in the predicted values.

**Fig. 1.**
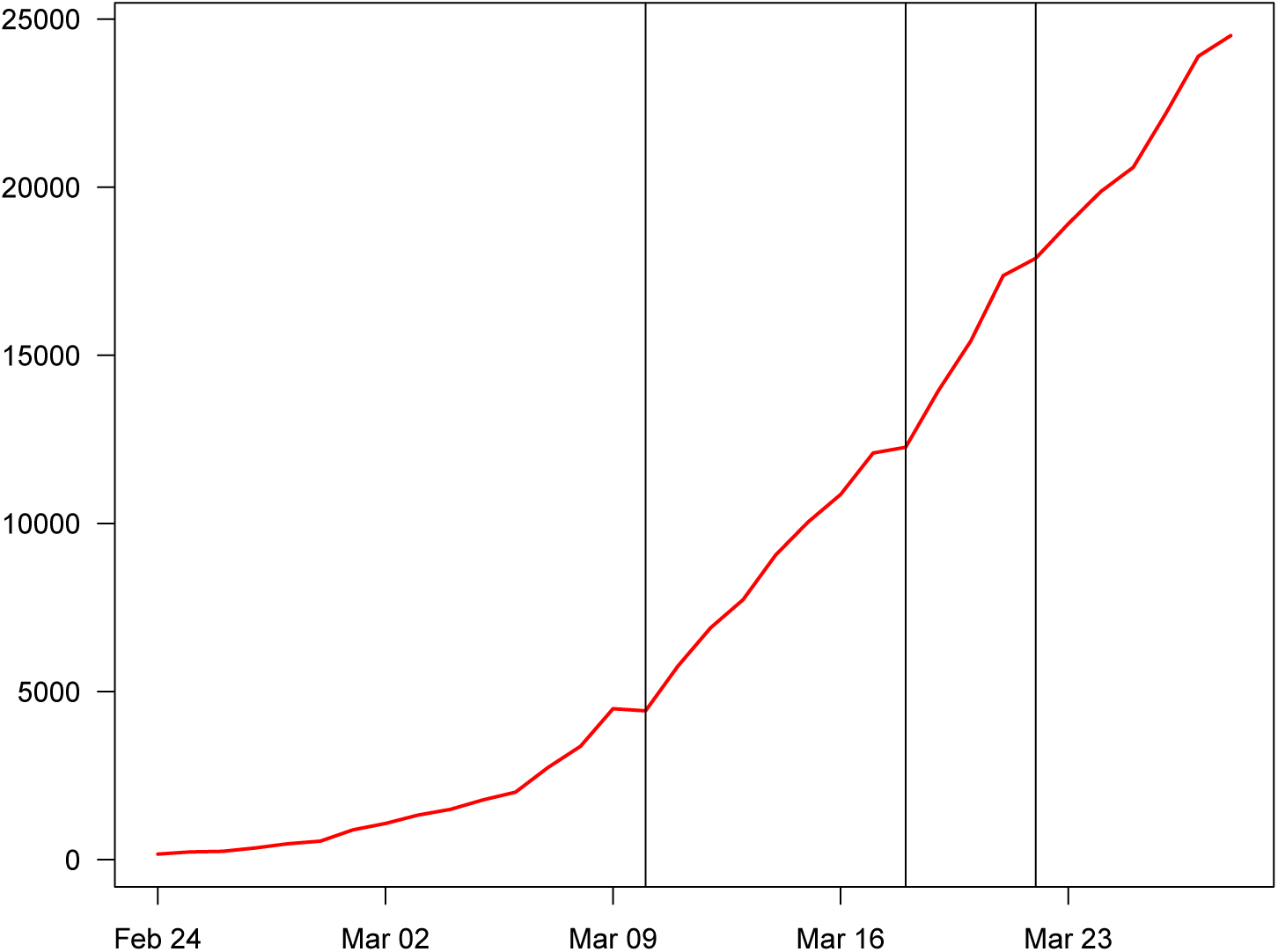
Number of people tested positive (Lombardia): original data.

In order to correct those data, a Kalman–smoother state–space model (Simon (2001)) has been applied. In particular, the Kalman smoother adopted is of the type *fixed point smoothing*. This algorithm is designed to obtain the estimate of a realization ŵ_*N*_ (the time *t*_*N*_ is fixed *N < K*) of a given random variable *W*_*t*_, given a set of observations ***Z***_*k*_ = *{****z***_*k*_|0 *≤ N ≤ k}* Sage and Melsa (1971).

In figure 2 the corrected version of the series ^1^***V***_1,*t*_ – resulting by applying the Kalman smoother – is depicted. Not only this series lends itself to a better visual inspection but, more importantly, is more suitable to be processed by the adopted prediction model.

**Fig. 2.**
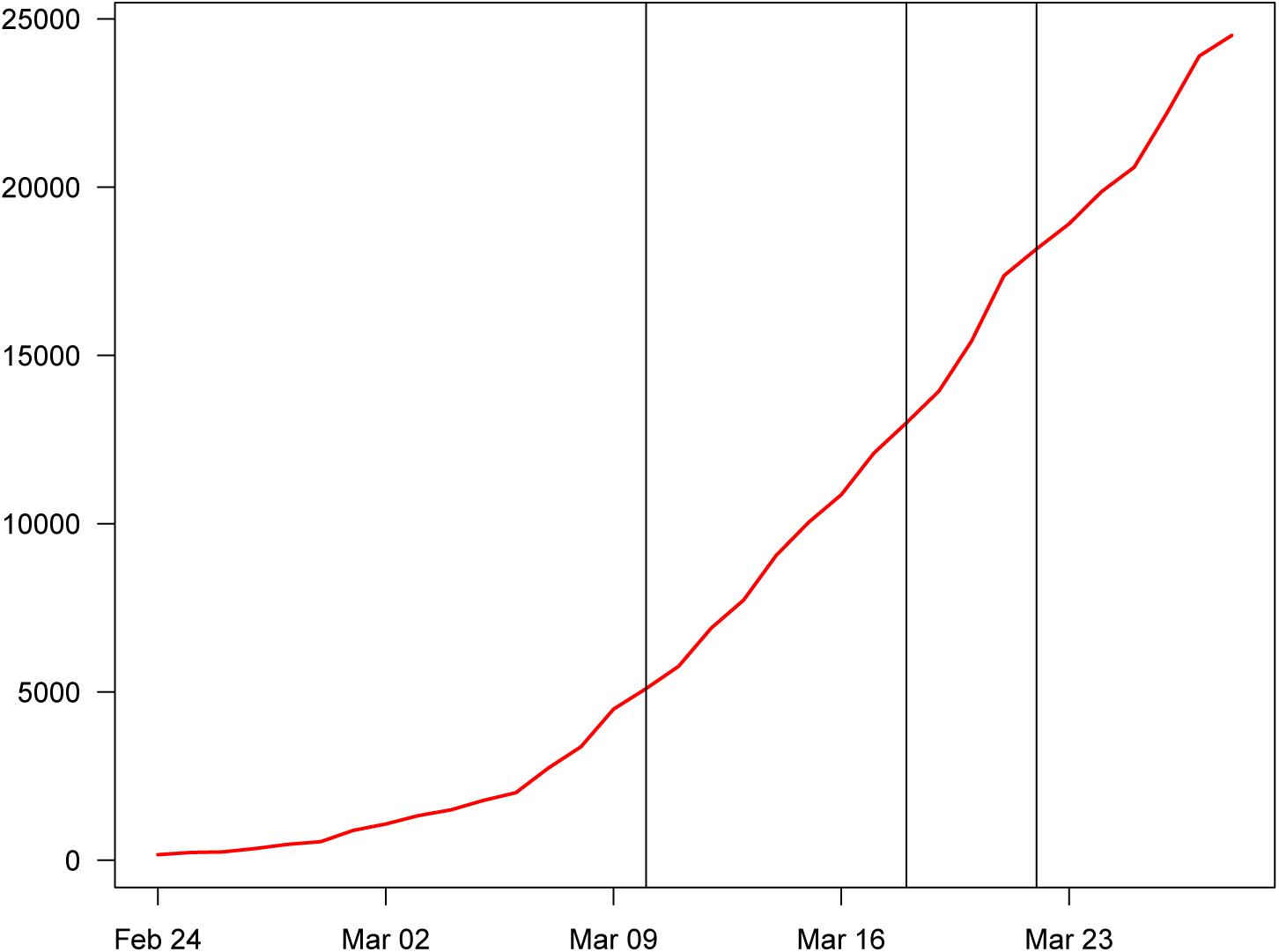
Number of people tested positive (Lombardia): data adjusted via Kalman filter.

## 4. Theoretical framework

The approach used in this paper relies on *i*) the theory of stochastic process and *ii*) a resampling method. While the former is necessary to generate the input (predicted values) of the bootstrap algorithm, as well as to justify the employment of the outlier correction method, the latter serves the purpose of

1. generating the final predictions, which are affected by a reduced amount of uncertainty (with respect to those generated by the stochastic model)
2. yielding the related confidence intervals.

### 4.1. The stochastic processes paradigm

The approach proposed in the present paper relies on the assumption that the (transformed) time series ^*K*^***V*** *j, t* and ^*K*^***V*** *j, t* are approximately a realization of a process of the type ARMA (Autoregressive Moving Average) (Makridakis and Hibon (1997)).

Let *X* = (*X*_*t*_)_*t∈*ℤ_ be a real 2^*nd*^ order stationary process, it is said to admit a ARMA(p,q) representation (p,q *∈* ℤ) if, for some constant *a*_1_….*a*_*p*_,*b*_1_…*b*_*q*_, will be:

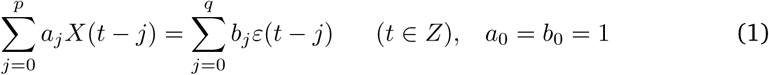

under the following conditions:

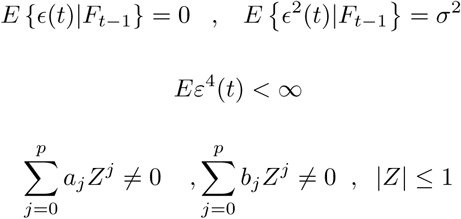

Here *F*_*t*_ denotes the sigma algebra induced by ϵ (*j*), *j ≤ t* and 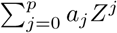 and 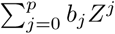 are assumed not to have common zero.

The above conditions assure that *X*_*t*_ can be represented as:

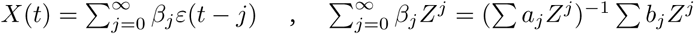

with *β*_*j*_ decreasing to 0 at geometric rate.

The dynamics of the series under investigations are not suitable for this theoretical framework which requires 2^*nd*^ – order stationarity; this is achieved by pre-processing the series according to the following filter: log(∇^*d*^), being the symbol ∇ the difference operator and the exponent *d* indicating the order of the difference. To fully understand the role played by ∇, the backward operator *B* is now introduced. In essence, *B* moves the time index of an observation back by *p* time intervals, i.e. *B*^*p*^*x*_*t*_ = *X*_*t−p*_, and thus we have that

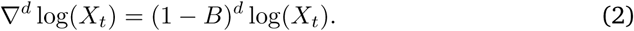

#### 4.2. The Resampling Method

In order to extract valuable information from our data and, at the same time, decrease the total amount of uncertainty associated to the outcomes of the ARMA model, a resampling procedure has been employed. Among the several resampling methods for dependent data available – many of which freely and publicly available in the form of powerful routines working under software packages such as Python^®^ or R^®^ – the adopted resampling method is of the type Maximum Entropy Bootstrap (*MEB*). Proposed by Vinod (2006) and subsequently improved (see, e.g., Vinod (2016)), it is based on basic assumptions which are different from those usually followed by standard schemes. In more details, while in the classic bootstrap an ensemble **Ω** represents the population of reference the observed time series is drawn from, in *MEB* a large number of ensembles (subsets), say *{****ω***_1_, …, ***ω***_*N*_ *}* becomes the elements belonging to **Ω**, each of them containing a large number of replicates *{x*_1_, …, *x*_*J*_ *}*.

Unlike standard bootstrap schemes, in the *MEB* case the resample set **Ω** mimics the observed realization of the underlying stochastic process, in *MEB* a large number of subsets, say *{****ω***_1_, …, ***ω***_*N*_*}* becomes the elements belonging to **Ω**, each of them containing a large number of replicates *{x*_1_, …, *x*_*J*_ *}*. Among the important features of the *MEB* scheme, it is worth mentioning the consistency of its bootstrap samples with the ergodic theorem (see, e.g., Birkhoff (1931)) and with the probabilistic structure of the observed time series. In Figure 3 an example of the application of *MEB* for the variable ^1^***V***_*t*,1_ is given.

**Fig. 3.**
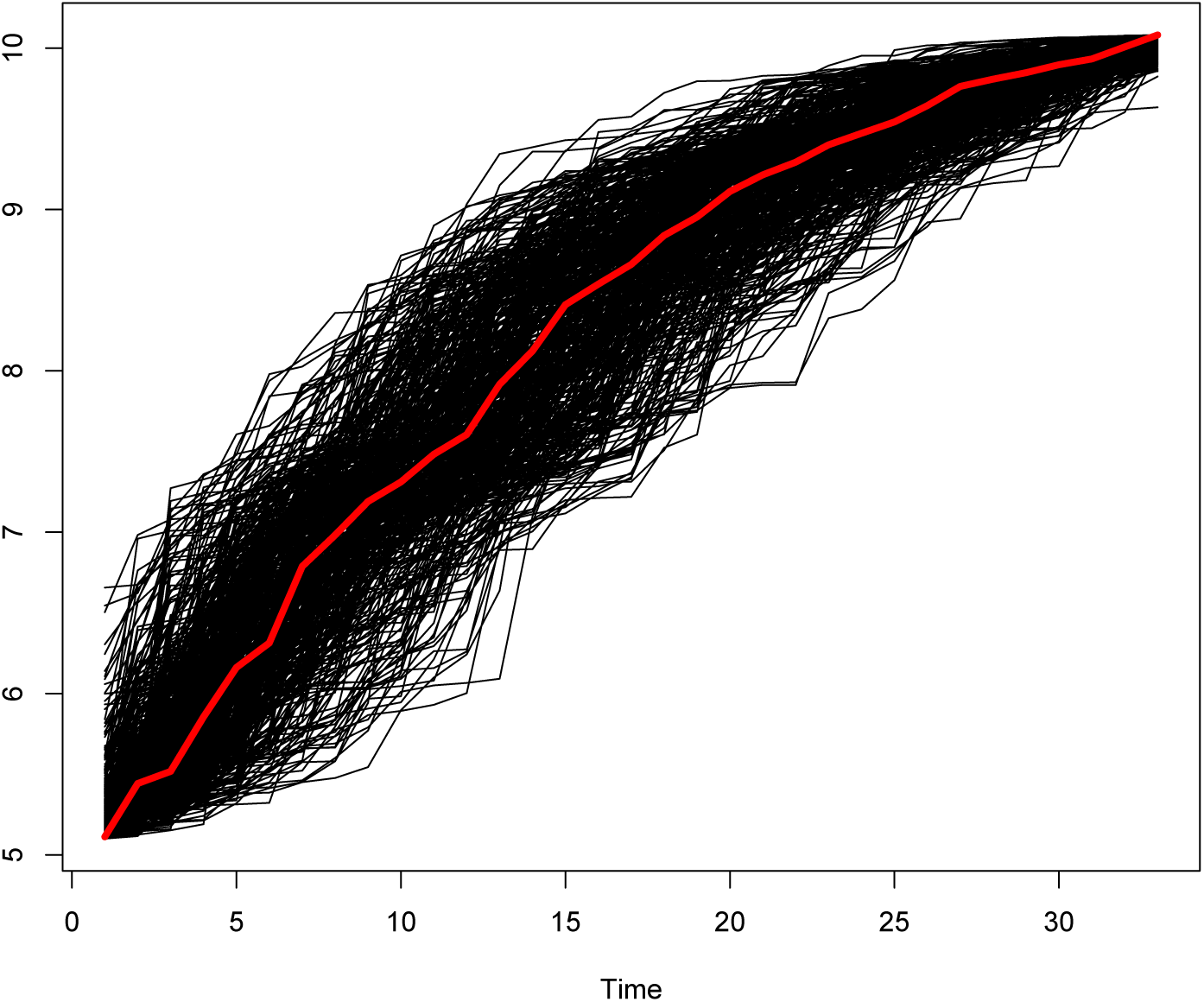
Lombardia: B= 500 Bootstrap replications performed via the *MEB* algorithm on the adjusted, log–transformed, data (in red the original time series is dpicted)

## 5. The forecasting method

In what follows, the proposed procedure is presented in a step-by-step fashion.

1. Eqn. 1 is estimated for both ***V***_*t*_ and ***U***_*t*_ so that the model orders (Eqn. 1) *M*_1_ and *M*_2_ become available;
2. for each time series ***V***_*t*_ and ***U***_*t*_ the *MEB* procedure is applied so that the sets *𝒱* and *𝒰* – each containing *B* = 500 “bona fide” replications – are available, i.e. 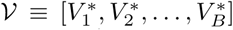 and 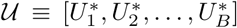 (in Figure 4 the set 𝒱 for the variable ^1^***V***_*t*,1_ is given);
3. for each of the replications stored in *𝒱*, Eqn. 1 is estimated according to the model order selected, i.e. *M*_1_, and the 1 to 10–step–ahead predictions – as well as the 5% and 95% bootstrap confidence interval – are generated;
4. the B predictions and the confidence intervals obtained in the previous step are stored in the *B×*3 matrix [***F***_*𝒱*_ (*h*); *h* = 1, 2, …, 10], whose columns are: lower bootstrap confidence interval, bootstrap prediction and upper bootstrap confidence interval, respectively denoted by the symbols 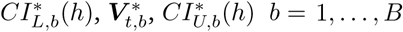.
5. the median value 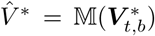 is then extracted along with the *≈* 95% confidence intervals 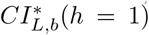and 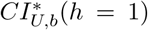, computed according to the t–percentile method. The explanation of this procedure goes beyond the scope of this paper, therefore the interested reader is referred to the excellent paper by Berkowitz and Kilian (2000).
6. 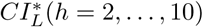 and 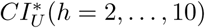 (the subscript *b* is omitted for brevity) are computed conditional to a subset of *𝒱*, say 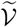, made up of the bootstrap replications whose range falls between the minimum and maximum values of the values of the confidence intervals computed for *h* = 1. In symbols:

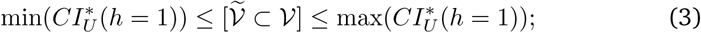
7. steps 1–6 are repeated for ***U***_*t*_, so that a new matrix of prediction of dimension *B ×* 3 is built, i.e. [***F***_*𝒱*_ (*h*) *h* = 1, 2, …, 10], whose columns are as in ***F***_*𝒰*_ (*h*) and denoted by the symbols 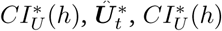.

**Fig. 4.**
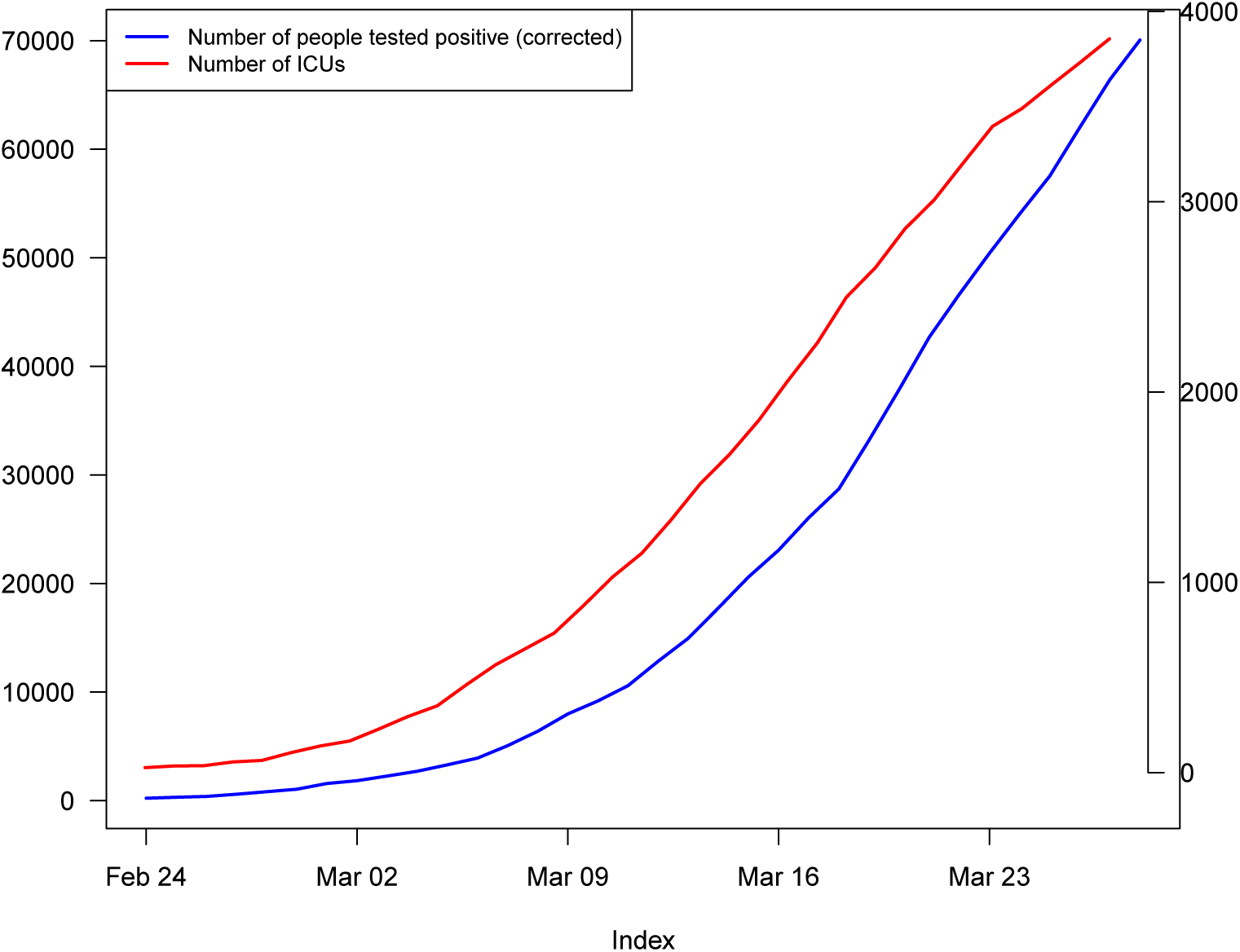
Italy, time series data of positive (data corrected via Kalman filter, left side axis) and of the number of people hospitalized in ICUs (right side axis)

Unfortunately, the whole procedure cannot be considered fully automatic since the estimation of Eqn. 1 (step 1) is required.

### 5.1. The adopted models

The stochastic model structures identified for both ***V***_*t*_ and ***U***_*t*_ are almost always of the type ARMA (1,0), with the exception of Campania (ARMA(0,1), for both the variables ***V***_*t*_ and ***U***_*t*_)) and Emilia Romagna, for which the best model for the variable ***U***_*t*_ is of the type ARMA(1,1). The most suitable prefilter (Eqn. 2) has been always of the type d=3 difference of the natural log of the variables of interest.

## 6. Empirical evidences

At the national level (data have been plotted in Figure 4) the peak in the number of CoViD-19–positives will be reached on 2 April, with a number of predicted positive close to 77,000. The maximum forecasted value for the occupied ICU – expected for April 4 – will be 4280. These values have been calculated using an indirect methodology, i.e. by summing up the estimates obtained at a disaggregated level. Regarding the results obtained at a disaggregated level, the models outcomes are now commented.

- Lombardia – the most affected region – will reach the peak of positive cases (25963) and of the demand of ICUs (1425) respectively on 2 and 4 April;
- Emilia Romagna is the second most affected Region by COVID-19 but still shows a very high number of victims. The trend of infected people will reach its peak on April 5th whereas the number of cases in Intensive Care will continue to grow at a progressively slower rate over the forecasting period;
- Veneto is the third Region for number of deaths. Here, the number of positive cases, as well as the number of cases in ICU, will reach the peak on April 3rd;
- for Piedmont – the fourth Region for number of victims – the predicted positive cases will reach the peak on March 29th (6635) whereas the persons in ICU will be 431 on 31 March, when the peak is predicted;
- Liguria will begin a process of relative reduction of positive cases as early as March 29th. The number of cases in Intensive Care, after a period of stability (lasting until 31 March), will start a slow decreasing path;
- Positive cases in Trentino Alto Adige – which incorporates the cities of Trento and Bolzano – are projected to be 2158 on March 30th and then a decreasing trend is expected. The ICU beds occupied in this region will reach its peak on around 3 April;
- The positive cases in Friuli Venezia Giulia show a relatively stable trend in the first half of the prediction interval with a peak around April 4th, after that the absolute number of cases will start decreasing. The number of cases in ICU will reach the peak between 30 March and 1 April;
- Valle D’Aosta is a small region which has been relatively less impacted by the virus. Here, a downward trend is expected to start on March 31st (for the positive cases) and around 31 March (cases in ICUs);
- The upward trend in the number of positive cases of Lazio is estimated to stop on 31 March and to reach the minimum at the end of the forecasting people (1821 cases). The number of ICU cases is estimated to reach its peak on the period 1–3 April;
- The Macro-area Center will reach its peak at the very beginning of the month of April (for the variable ***V***) whereas for the variable ***U*** the estimated peak day is around 31 March;
- Campania will reach the peak of contagions on April 5 whereas ICU cases will do on the previous day;
- The remaining southern regions (Abruzzo, Molise, Puglia, Basilicata, Calabria, Sicily and Sardinia) will show an upward trend in the number of future positive cases lasting until 6 April, where 6355 cases are predicted. The number of persons requiring an ICU will reach the peak on 4 April (348 is the estimated number of cases).

**Table 1.** 10–step–ahead predictions for the variable ***V***_*t*_, number of persons tested positive

| Italy |  |  |  |
| --- | --- | --- | --- |
| | $CI_L^*(h)$ | $\hat{V}_t^*$ | $CI_U^*(h)$ |
| 29 March | 66504 | 71586 | 76647 |
| 30 March | 69753 | 73409 | 79373 |
| 31 March | 70760 | 75696 | 82773 |
| 1 April | 70817 | 76723 | 86430 |
| 2 April | 69925 | 77811 | 90800 |
| 3 April | 69309 | 76952 | 96806 |
| 4 April | 68480 | 75927 | 102946 |
| 5 April | 66531 | 74860 | 109265 |
| 6 April | 64940 | 75006 | 117550 |
| 7 April | 60197 | 74875 | 126205 |
| Lombardia |  |  |  |
| | $CI_L^*(h)$ | $\hat{V}_t^*$ | $CI_U^*(h)$ |
| 29 March | 22538 | 25214 | 26384 |
| 30 March | 23296 | 25456 | 26790 |
| 31 March | 23969 | 25792 | 27565 |
| 1 April | 23864 | 25842 | 27634 |
| 2 April | 23662 | 25963 | 28230 |
| 3 April | 23209 | 25675 | 28342 |
| 4 April | 22709 | 25717 | 28781 |
| 5 April | 21964 | 25067 | 29524 |
| 6 April | 20802 | 24431 | 30131 |
| 7 April | 19619 | 23752 | 30628 |
| Piedmont |  |  |  |
| | $CI_L^*(h)$ | $\hat{V}_t^*$ | $CI_U^*(h)$ |
| 29 March | 6115 | 6635 | 7017 |
| 30 March | 6264 | 6610 | 7193 |
| 31 March | 6209 | 6568 | 7481 |
| 1 April | 6079 | 6401 | 7942 |
| 2 April | 5837 | 6152 | 8271 |
| 3 April | 5502 | 5898 | 8590 |
| 4 April | 5134 | 5566 | 8753 |
| 5 April | 4666 | 5135 | 8995 |
| 6 April | 4178 | 4732 | 9279 |
| 7 April | 3707 | 4289 | 9417 |

Liguria
| | $CI_L^*(h)$ | $\hat{V}_t^*$ | $CI_U^*(h)$ |
| --- | --- | --- | --- |
| 29 March | 2026 | 2062 | 2217 |
| 30 March | 2007 | 2048 | 2242 |
| 31 March | 1955 | 2019 | 2361 |
| 1 April | 1891 | 1969 | 2416 |
| 2 April | 1804 | 1900 | 2515 |
| 3 April | 1701 | 1805 | 2580 |
| 4 April | 1589 | 1709 | 2645 |
| 5 April | 1470 | 1590 | 2748 |
| 6 April | 1344 | 1440 | 2917 |
| 7 April | 1214 | 1311 | 3090 |

Valle D'Aosta
| | $CI_L^*(h)$ | $\hat{V}_t^*$ | $CI_U^*(h)$ |
| --- | --- | --- | --- |
| 29 March | 404 | 480 | 512 |
| 30 March | 410 | 491 | 524 |
| 31 March | 417 | 501 | 546 |
| 1 April | 420 | 500 | 551 |
| 2 April | 411 | 494 | 551 |
| 3 April | 382 | 483 | 542 |
| 4 April | 359 | 462 | 526 |
| 5 April | 339 | 443 | 541 |
| 6 April | 301 | 412 | 530 |
| 7 April | 267 | 383 | 516 |

Veneto
| | $CI_L^*(h)$ | $\hat{V}_t^*$ | $CI_U^*(h)$ |
| --- | --- | --- | --- |
| 29 March | 6346 | 7145 | 7559 |
| 30 March | 6569 | 7213 | 7749 |
| 31 March | 6706 | 7431 | 8067 |
| 1 April | 6721 | 7463 | 8144 |
| 2 April | 6535 | 7539 | 8415 |
| 3 April | 6414 | 7469 | 8758 |
| 4 April | 6073 | 7374 | 9168 |
| 5 April | 5703 | 7182 | 9214 |
| 6 April | 5303 | 6842 | 9593 |
| 7 April | 4798 | 6537 | 10203 |

Friuli Venezia Giulia Infetti
| | $CI_L^*(h)$ | $\hat{V}_t^*$ | $CI_U^*(h)$ |
| --- | --- | --- | --- |
| 29 March | 1029 | 1124 | 1167 |
| 30 March | 1083 | 1138 | 1195 |
| 31 March | 1100 | 1169 | 1226 |
| 1 April | 1104 | 1189 | 1269 |
| 2 April | 1109 | 1212 | 1267 |
| 3 April | 1097 | 1209 | 1272 |
| 4 April | 1060 | 1230 | 1287 |
| 5 April | 1035 | 1203 | 1305 |
| 6 April | 1005 | 1202 | 1333 |
| 7 April | 976 | 1208 | 1343 |

Emilia Romagna
| | $CI_L^*(h)$ | $\hat{V}_t^*$ | $CI_U^*(h)$ |
| --- | --- | --- | --- |
| 29 March | 9163 | 10500 | 11265 |
| 30 March | 9464 | 10975 | 11717 |
| 31 March | 9535 | 11360 | 12176 |
| 1 April | 9587 | 11613 | 12562 |
| 2 April | 9476 | 11815 | 12954 |
| 3 April | 9276 | 11943 | 13291 |
| 4 April | 9002 | 12132 | 13539 |
| 5 April | 8528 | 12244 | 13720 |
| 6 April | 8034 | 12125 | 13995 |
| 7 April | 7311 | 11966 | 14189.9250136506 |

Trentino Alto Adige
| | $CI_L^*(h)$ | $\hat{V}_t^*$ | $CI_U^*(h)$ |
| --- | --- | --- | --- |
| 29 March | 2021 | 2147 | 2324 |
| 30 March | 2076 | 2158 | 2488 |
| 31 March | 2114 | 2124 | 2617 |
| 1 April | 2110 | 2083 | 2707 |
| 2 April | 2097 | 1988 | 2787 |
| 3 April | 2064 | 1868 | 2764 |
| 4 April | 2023 | 1725 | 2742 |
| 5 April | 1944 | 1548 | 2751 |
| 6 April | 1851 | 1373 | 2718 |
| 7 April | 1707 | 1200 | 2641 |

Lazio
| | $CI_L^*(h)$ | $\hat{V}_t^*$ | $CI_U^*(h)$ |
| --- | --- | --- | --- |
| 29 March | 2013 | 2208 | 2355 |
| 30 March | 2160 | 2244 | 2436 |
| 31 March | 2182 | 2298 | 2553 |
| 1 April | 2153 | 2279 | 2666 |
| 2 April | 2161 | 2256 | 2785 |
| 3 April | 2140 | 2222 | 2991 |
| 4 April | 2100 | 2154 | 3132 |
| 5 April | 2041 | 2046 | 3233 |
| 6 April | 1908 | 1956 | 3485 |
| 7 April | 1768 | 1821 | 3782 |

Macro Area Center
| | $CI_L^*(h)$ | $\hat{V}_t^*$ | $CI_U^*(h)$ |
| --- | --- | --- | --- |
| 29 March | 6822 | 7499 | 7911 |
| 30 March | 7104 | 7671 | 8087 |
| 31 March | 7130 | 7864 | 8396 |
| 1 April | 7001 | 7896 | 8741 |
| 2 April | 6758 | 7889 | 8987 |
| 3 April | 6424 | 7851 | 9156 |
| 4 April | 6078 | 7813 | 9695 |
| 5 April | 5512 | 7718 | 10081 |
| 6 April | 4925 | 7494 | 10580 |
| 7 April | 4397 | 7238 | 10941 |

Campania
| | $CI_L^*(h)$ | $\hat{V}_t^*$ | $CI_U^*(h)$ |
| --- | --- | --- | --- |
| 29 March | 1169 | 1453 | 1549 |
| 30 March | 1267 | 1494 | 1662 |
| 31 March | 1293 | 1527 | 1747 |
| 1 April | 1287 | 1548 | 1835 |
| 2 April | 1280 | 1582 | 1898 |
| 3 April | 1252 | 1619 | 1953 |
| 4 April | 1222 | 1640 | 1972 |
| 5 April | 1160 | 1649 | 2020 |
| 6 April | 1090 | 1648 | 2063 |
| 7 April | 994 | 1645 | 2145 |

Macro Area Sud
| | $CI_L^*(h)$ | $\hat{V}_t^*$ | $CI_U^*(h)$ |
| --- | --- | --- | --- |
| 29 March | 4380 | 5226 | 5662 |
| 30 March | 4646 | 5406 | 5846 |
| 31 March | 4689 | 5648 | 6246 |
| 1 April | 4705 | 5791 | 6528 |
| 2 April | 4496 | 5931 | 6902 |
| 3 April | 4122 | 6129 | 7325 |
| 4 April | 3841 | 6169 | 7695 |
| 5 April | 3445 | 6346 | 8141 |
| 6 April | 2981 | 6355 | 8787 |
| 7 April | 2512 | 6345 | 9397 |

**Table 2.**
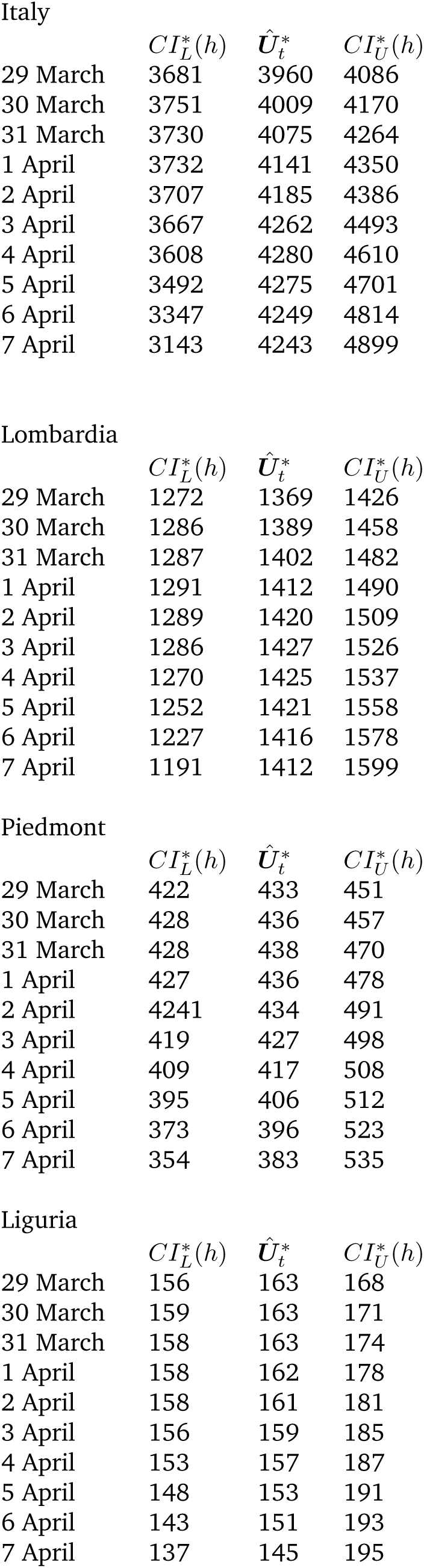
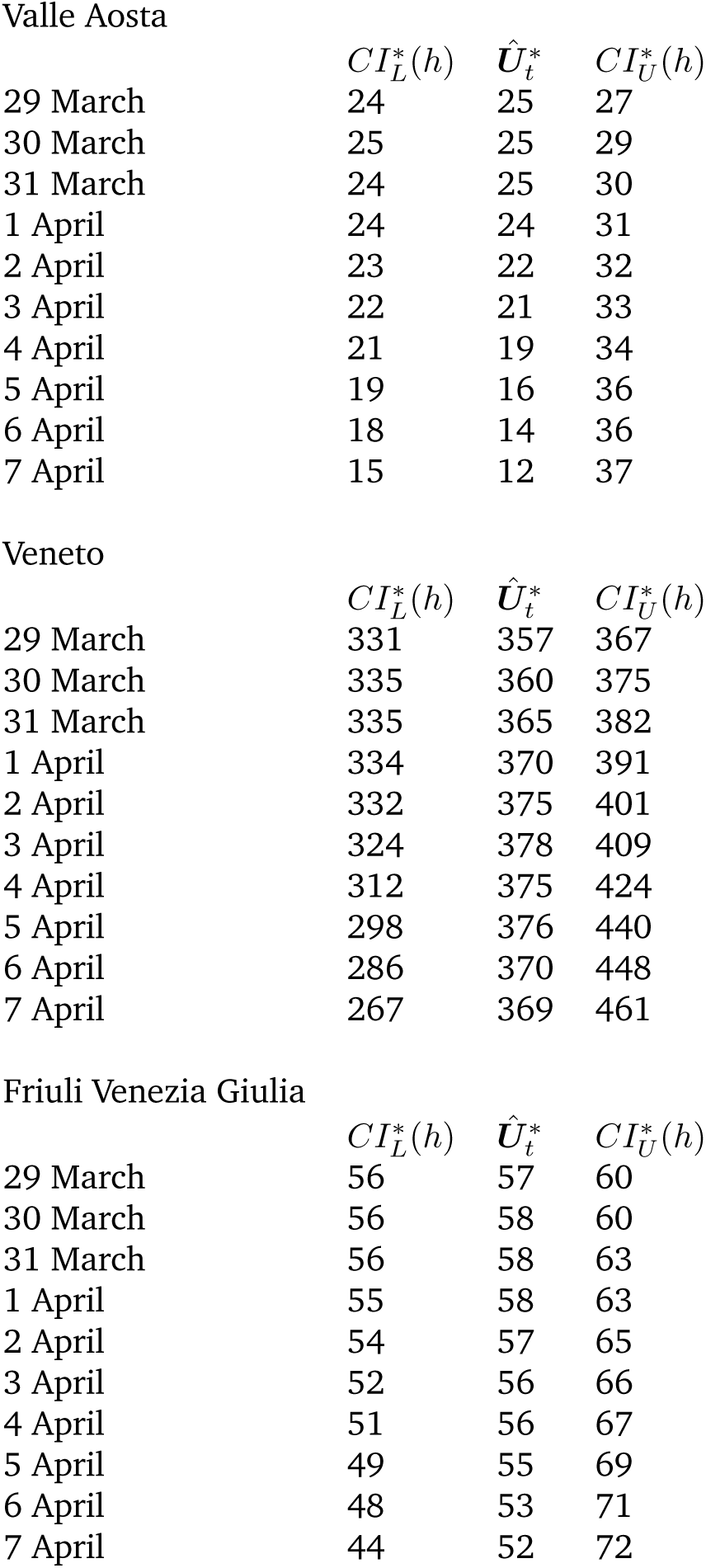
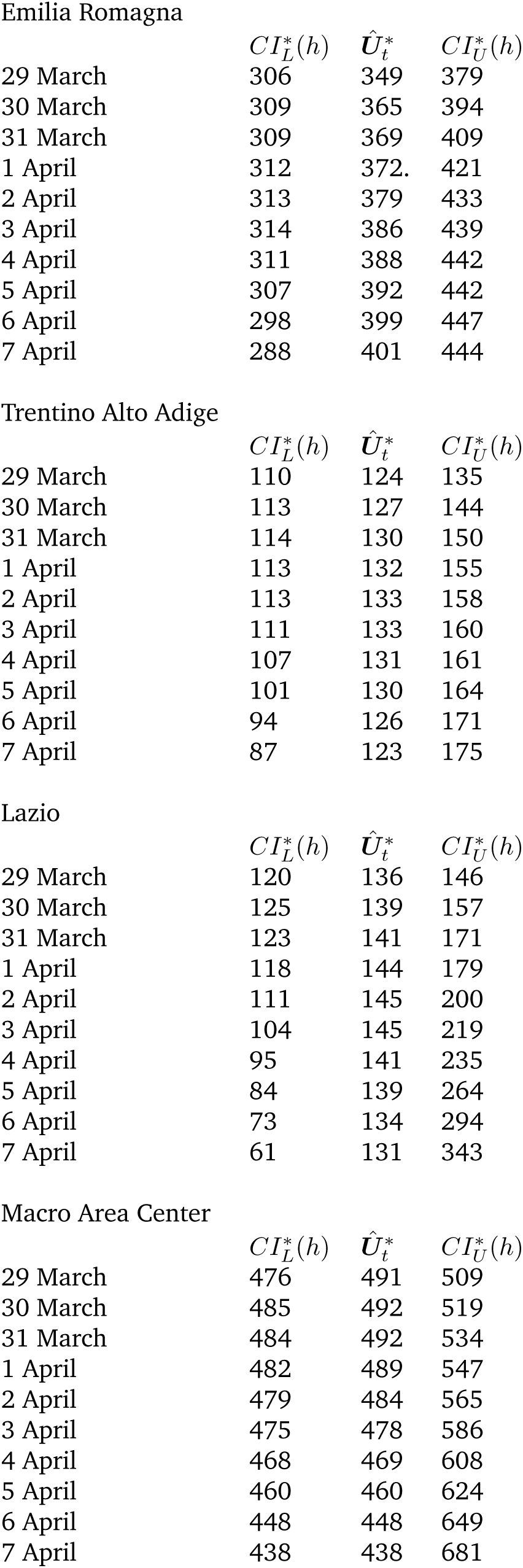
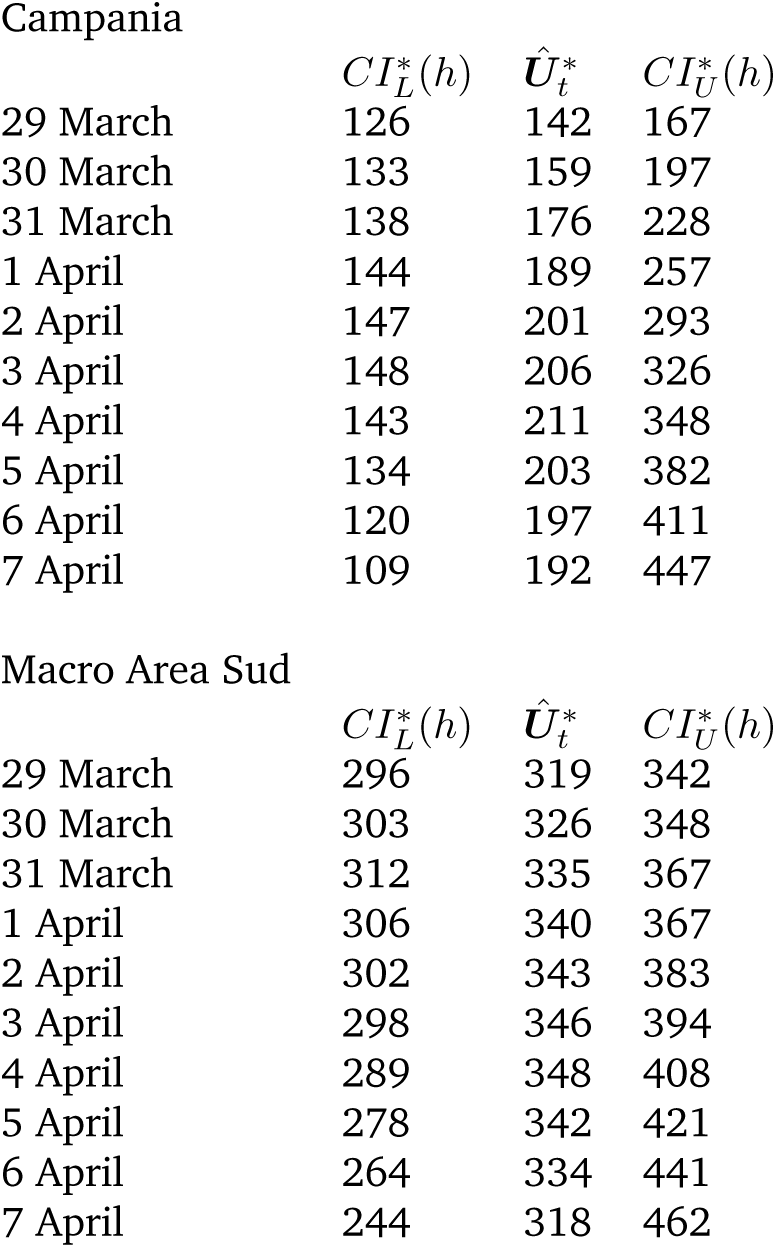
10–step–ahead predictions for the variable ***U***_*t*_ (people in ICUs)

| Italy |  |  |  |
| --- | --- | --- | --- |
| | $CI_L^*(h)$ | $\hat{U}_t^*$ | $CI_U^*(h)$ |
| 29 March | 3681 | 3960 | 4086 |
| 30 March | 3751 | 4009 | 4170 |
| 31 March | 3730 | 4075 | 4264 |
| 1 April | 3732 | 4141 | 4350 |
| 2 April | 3707 | 4185 | 4386 |
| 3 April | 3667 | 4262 | 4493 |
| 4 April | 3608 | 4280 | 4610 |
| 5 April | 3492 | 4275 | 4701 |
| 6 April | 3347 | 4249 | 4814 |
| 7 April | 3143 | 4243 | 4899 |
| Lombardia |  |  |  |
| | $CI_L^*(h)$ | $\hat{U}_t^*$ | $CI_U^*(h)$ |
| 29 March | 1272 | 1369 | 1426 |
| 30 March | 1286 | 1389 | 1458 |
| 31 March | 1287 | 1402 | 1482 |
| 1 April | 1291 | 1412 | 1490 |
| 2 April | 1289 | 1420 | 1509 |
| 3 April | 1286 | 1427 | 1526 |
| 4 April | 1270 | 1425 | 1537 |
| 5 April | 1252 | 1421 | 1558 |
| 6 April | 1227 | 1416 | 1578 |
| 7 April | 1191 | 1412 | 1599 |
| Piedmont |  |  |  |
| | $CI_L^*(h)$ | $\hat{U}_t^*$ | $CI_U^*(h)$ |
| 29 March | 422 | 433 | 451 |
| 30 March | 428 | 436 | 457 |
| 31 March | 428 | 438 | 470 |
| 1 April | 427 | 436 | 478 |
| 2 April | 4241 | 434 | 491 |
| 3 April | 419 | 427 | 498 |
| 4 April | 409 | 417 | 508 |
| 5 April | 395 | 406 | 512 |
| 6 April | 373 | 396 | 523 |
| 7 April | 354 | 383 | 535 |
| Liguria |  |  |  |
| | $CI_L^*(h)$ | $\hat{U}_t^*$ | $CI_U^*(h)$ |
| 29 March | 156 | 163 | 168 |
| 30 March | 159 | 163 | 171 |
| 31 March | 158 | 163 | 174 |
| 1 April | 158 | 162 | 178 |
| 2 April | 158 | 161 | 181 |
| 3 April | 156 | 159 | 185 |
| 4 April | 153 | 157 | 187 |
| 5 April | 148 | 153 | 191 |
| 6 April | 143 | 151 | 193 |
| 7 April | 137 | 145 | 195 |

Valle Aosta
| | $CI_L^*(h)$ | $\hat{U}_t^*$ | $CI_U^*(h)$ |
| --- | --- | --- | --- |
| 29 March | 24 | 25 | 27 |
| 30 March | 25 | 25 | 29 |
| 31 March | 24 | 25 | 30 |
| 1 April | 24 | 24 | 31 |
| 2 April | 23 | 22 | 32 |
| 3 April | 22 | 21 | 33 |
| 4 April | 21 | 19 | 34 |
| 5 April | 19 | 16 | 36 |
| 6 April | 18 | 14 | 36 |
| 7 April | 15 | 12 | 37 |

Veneto
| | $CI_L^*(h)$ | $\hat{U}_t^*$ | $CI_U^*(h)$ |
| --- | --- | --- | --- |
| 29 March | 331 | 357 | 367 |
| 30 March | 335 | 360 | 375 |
| 31 March | 335 | 365 | 382 |
| 1 April | 334 | 370 | 391 |
| 2 April | 332 | 375 | 401 |
| 3 April | 324 | 378 | 409 |
| 4 April | 312 | 375 | 424 |
| 5 April | 298 | 376 | 440 |
| 6 April | 286 | 370 | 448 |
| 7 April | 267 | 369 | 461 |

Friuli Venezia Giulia
| | $CI_L^*(h)$ | $\hat{U}_t^*$ | $CI_U^*(h)$ |
| --- | --- | --- | --- |
| 29 March | 56 | 57 | 60 |
| 30 March | 56 | 58 | 60 |
| 31 March | 56 | 58 | 63 |
| 1 April | 55 | 58 | 63 |
| 2 April | 54 | 57 | 65 |
| 3 April | 52 | 56 | 66 |
| 4 April | 51 | 56 | 67 |
| 5 April | 49 | 55 | 69 |
| 6 April | 48 | 53 | 71 |
| 7 April | 44 | 52 | 72 |

Emilia Romagna
| | $CI_L^*(h)$ | $\hat{U}_t^*$ | $CI_U^*(h)$ |
| --- | --- | --- | --- |
| 29 March | 306 | 349 | 379 |
| 30 March | 309 | 365 | 394 |
| 31 March | 309 | 369 | 409 |
| 1 April | 312 | 372. | 421 |
| 2 April | 313 | 379 | 433 |
| 3 April | 314 | 386 | 439 |
| 4 April | 311 | 388 | 442 |
| 5 April | 307 | 392 | 442 |
| 6 April | 298 | 399 | 447 |
| 7 April | 288 | 401 | 444 |

Trentino Alto Adige
| | $CI_L^*(h)$ | $\hat{U}_t^*$ | $CI_U^*(h)$ |
| --- | --- | --- | --- |
| 29 March | 110 | 124 | 135 |
| 30 March | 113 | 127 | 144 |
| 31 March | 114 | 130 | 150 |
| 1 April | 113 | 132 | 155 |
| 2 April | 113 | 133 | 158 |
| 3 April | 111 | 133 | 160 |
| 4 April | 107 | 131 | 161 |
| 5 April | 101 | 130 | 164 |
| 6 April | 94 | 126 | 171 |
| 7 April | 87 | 123 | 175 |

Lazio
| | $CI_L^*(h)$ | $\hat{U}_t^*$ | $CI_U^*(h)$ |
| --- | --- | --- | --- |
| 29 March | 120 | 136 | 146 |
| 30 March | 125 | 139 | 157 |
| 31 March | 123 | 141 | 171 |
| 1 April | 118 | 144 | 179 |
| 2 April | 111 | 145 | 200 |
| 3 April | 104 | 145 | 219 |
| 4 April | 95 | 141 | 235 |
| 5 April | 84 | 139 | 264 |
| 6 April | 73 | 134 | 294 |
| 7 April | 61 | 131 | 343 |

Macro Area Center
| | $CI_L^*(h)$ | $\hat{U}_t^*$ | $CI_U^*(h)$ |
| --- | --- | --- | --- |
| 29 March | 476 | 491 | 509 |
| 30 March | 485 | 492 | 519 |
| 31 March | 484 | 492 | 534 |
| 1 April | 482 | 489 | 547 |
| 2 April | 479 | 484 | 565 |
| 3 April | 475 | 478 | 586 |
| 4 April | 468 | 469 | 608 |
| 5 April | 460 | 460 | 624 |
| 6 April | 448 | 448 | 649 |
| 7 April | 438 | 438 | 681 |

Campania
| | $CI_L^*(h)$ | $\hat{U}_t^*$ | $CI_U^*(h)$ |
| --- | --- | --- | --- |
| 29 March | 126 | 142 | 167 |
| 30 March | 133 | 159 | 197 |
| 31 March | 138 | 176 | 228 |
| 1 April | 144 | 189 | 257 |
| 2 April | 147 | 201 | 293 |
| 3 April | 148 | 206 | 326 |
| 4 April | 143 | 211 | 348 |
| 5 April | 134 | 203 | 382 |
| 6 April | 120 | 197 | 411 |
| 7 April | 109 | 192 | 447 |

Macro Area Sud
| | $CI_L^*(h)$ | $\hat{U}_t^*$ | $CI_U^*(h)$ |
| --- | --- | --- | --- |
| 29 March | 296 | 319 | 342 |
| 30 March | 303 | 326 | 348 |
| 31 March | 312 | 335 | 367 |
| 1 April | 306 | 340 | 367 |
| 2 April | 302 | 343 | 383 |
| 3 April | 298 | 346 | 394 |
| 4 April | 289 | 348 | 408 |
| 5 April | 278 | 342 | 421 |
| 6 April | 264 | 334 | 441 |
| 7 April | 244 | 318 | 462 |

## 7. Disclaimer

The views and opinions expressed in this article are those of the author and do not necessarily reflect the official policy or position of the Italian National Institute of Statistics.

## Data Availability

All the data used in my paper can be freely accessed on the internet

https://www.iss.it/

